# Self-Directed Home-Based Dim-Light Melatonin Onset Collection: The Circadia Pilot Study

**DOI:** 10.1101/2023.05.26.23290467

**Authors:** Gregory Bormes, Jessica Love, Akeju Oluwaseun, Jakob Cherry, Lovemore Kunorozva, Salim Qadri, Shadab A. Rahman, Brandon Westover, John Winkelman, Jacqueline M. Lane

## Abstract

**Study Objectives:** To test the feasibility of a novel at-home salivary Dim Light Melatonin Onset (DLMO) assessment protocol to measure the endogenous circadian phase of 10 individuals (1 Advanced Sleep-Wake Phase Disorder patient (ASWPD), 4 Delayed Sleep-Wake Phase Disorder patients (DSWPD), and 5 controls).

**Methods:** **The study involved 10 participants (sex at birth: females = 9; male= 1), who ranged between 27 to 63 years old, with an average age of 38 years old. Our study population consisted of 7 individuals who identified as white and 3 who identified as Asian. Our participants were diverse in gender identity (woman = 7, male = 1, transgender = 1, nonbinary = 1, none = 1)**.The study tracked the sleep and activity patterns of 10 individuals over a 5-6 week period using self-reported online sleep diaries and objective actigraphy data. Participants completed two self-directed DLMO assessments, approximately one week apart, adhering to objective compliance measures. Participants completed the study entirely remotely: they completed all sleep diaries and other evaluations online and were mailed a kit with all materials needed to perform the actigraphy and at-home sample collections.

**Results:** Salivary DLMO times were calculated for 8/10 participants using the Hockeystick method. DLMO times were on average 3 hours and 18 minutes earlier than self-reported sleep onset times (DSPD: 12:04 AM, controls: 9:55 PM.) Among the 6 participants for whom we calculated two separate DLMO times, DLMOs 1 and 2 were 96% correlated (p<0.0005.)

**Conclusions:** Our results indicate that self-directed, at-home DLMO assessments are feasible and accurate. The current protocol may serve as a framework to reliably assess circadian phase in both clinical and general populations.

## Introduction

Circadian rhythm sleep-wake disorders (CRSWD) describe a type of rare disorder involving a misalignment of internal circadian rhythms and the external environment that can stem from a variety of social, environmental, and genetic factors.^1,2,3,4^ Two CRSWDs in particular, Advanced Sleep-Wake Phase Disorder (ASWPD) and Delayed Sleep-Wake Phase Disorder (DSWPD have been shown to have a strong genetic component.^5,6^ Due to shifted physiological processes relative to the external environment, CRSWDs can lead to altered circadian timing which is frequently associated with neuropsychiatric^7^ and cardiometabolic comorbidities,^8^ as a risk factor for suicide,^9^ pharmaco-resistance^10,11^ or as a preclinical sign of Alzheimer’s of Parkinson’s disease.^12,13^ By the International Classification of Sleep Disorders (ICSD) criteria, ASWPD is prevalent in up to 0.21% of the population,^5^ and DSWPD in about 3%,^14,15,16^ although CRSWDs are often undiagnosed or misdiagnosed, making the true prevalence difficult to estimate.^17^

Diagnosis of a circadian rhythm disorder typically involves assessment of sleep timing while ruling out confounding factors. Most often, symptoms are captured by self-report in a sleep log and/or in concert with objectively measured sleep timing through actigraphy. These methods capture symptoms, but do not assess underlying physiology. The current gold standard method for estimating circadian phase is a Dim Light Melatonin Onset (DLMO).^18^ This often requires an in-person visit to a sleep or circadian clinic, which presents geographic, financial, and temporal barriers.^19^ Scheduling visits, traveling to clinics, and staying at a clinic overnight all add to participant burden.

Compounding these difficulties, wait times for treatment at circadian disorders sleep clinics are extremely long. DLMO assessments are not typically covered by insurance, which further complicates the use of DLMO testing for patients’ CRSWD diagnosis.^16^ More accessible methods of circadian phase assessment have been identified as a top priority in CRSWD research.^16^ With these barriers in mind, we designed the Circadia Study to be as accessible as possible.

Previously, Burgess et. al demonstrated that DLMOs measured at home and supervised by a trained technician correlate highly with DLMOs measured in a laboratory in a DSWPD population, indicating home-based DLMOs as a feasible diagnostic tool for CRSWDs.^20,21^ In the Circadia Pilot study, a direct-to-participant study, we developed a novel method for measuring DLMO at-home that can be completed by an individual independently. This remote, self-directed assessment method is less expensive than traditional DLMOs and not limited by geographic barriers, thus making circadian phase data much easier to obtain and easier to complete for patients. Additionally, using our methods, circadian phase data can more easily be incorporated into research on myriad conditions where circadian rhythms may play a role.^6,22^

In this pilot study, we aimed to test the feasibility of a novel at-home DLMO assessment design. We collected circadian phase and daily sleep data in 10 participants over a 4-6 week span. Each participant completed two DLMOs remotely, approximately one week apart, which we used to calculate DLMO times for each participant. We objectively measured compliance with study conditions for the at-home DLMOs, assessed congruence between repeated measures, and identified areas for improvement.

## Methods

### Recruitment

The study was advertised on the MassGeneralBrigham Rally recruitment site (https://rally.massgeneralbrigham.org/) and through online CRSWD patient advocacy groups. [Supplement Material: Rally Advertisement] Participants were recruited from September 2021 - May 2022. After individuals expressed interest in the study through Rally, we contacted them via email with detailed information and a link to sign up on our web portal. [Supplemental Material: Rally Follow-up Email.] After obtaining informed consent, we enrolled 5 individuals with a diagnosed CRSWD (1 with ASWPD and 4 with DSWPD), and 5 controls with no diagnosed CRSWD. [Table: Demographics.] Final follow-up occurred in July 2022. The study was approved by MassGeneral IRB (IRB Number: 2021P003666).

### Online Study Portal

We created a Circadia Study online study portal through collaboration with the Broad Institute Data Science Platform using their Pepper Platform (https://circadiastudy.org/.) Interested individuals were directed to the study portal where they could find more information about our study, including informational videos describing the study procedure [Supplemental Material: Instructional Videos.] On the portal, they completed a web questionnaire screener (n=11 questions) to automatically determine their eligibility. [Supplemental Material: Screening Questionnaire] Participants used the study portal to complete each online task throughout the study.

### Eligibility Criteria

Eligible participants included anyone over the age of 23^23^ who could read and write at an 8th-grade English reading level. Participants were excluded based on the following criteria: individuals under the age of 23; enrollment in school; current pregnancy or parturition within a year of study participation;^24^ current medications that may interfere with circulating melatonin levels; history of traumatic brain injury, stroke, or seizure disorder^25^; gingivitis, xerostomia, or periodontitis and blindness. Enrollment was limited to 5 controls with no diagnosed CRSWD and 5 individuals with a diagnosed CRSWD of Advanced or Delayed Sleep Wake Phase Disorder and symptoms present for longer than three months. [Supplemental Material: Eligibility Criteria Table.] Eligible participants consented and enrolled directly through the study portal.^19^

### Sleep Logging and DLMO Scheduling

Once a participant had enrolled, they landed on an introductory video that described the next steps of the study and directed them to begin sleep logging. Participants used our study website to begin sleep logging using a direct and secure connection of our custom portal to a sleep diary designed by researchers at Brigham and Women’s Hospital^26,27^ and self-schedule their two DLMO collection dates using Microsoft Bookings. We required the DLMOs to be approximately one week apart, but participants could otherwise choose any dates that worked best for their schedule. Participants logged their sleep online each day via the BWH Sleep Log using an online questionnaire (17 items) for the duration of their participation in the study (4-6 weeks.) [Supplemental Material: Sleep Log Questionnaire.] DLMO assessments were scheduled to begin 6 hours before their average bedtime estimated by study staff based on their online sleep diary data.^20^

### Circadia Study At-Home Kit

To ensure at-home kits were mailed to engaged participants only, after participants logged their sleep for five days, and scheduled their DLMO collections, we mailed them a Circadia Study kit complete with everything needed to complete their at-home DLMO. These kits included a personalized instruction manual [Supplemental Material: at-home Kit Instructions], an Actiwatch Spectrum Plus to record light and activity data, a light meter, blue light-blocking glasses, 2 sets of 9 salivettes in a closed plastic bottle, a time tracker lid for the bottles to record the exact time each sample was taken, two freezer bags and ice packs in which to store collected samples, and a prepaid shipping label to return the kit and samples. The kit contained miscellaneous extra materials that might be needed during the DLMO collections like toothbrushes, tea lights, labels, markers, extra salivettes and aluminum foil and contractor grade trash bags to darken any windows in the collection space [Supplemental Material: At-Home Kit Contents.] Further details on each kit content are found in the Supplemental Material: At-Home Kit Contents. In an unboxing video describing the contents of the kit, we instructed participants to begin wearing the Actiwatch on their non-dominant arm as soon as they received it. [Supplemental Material: Instructional Materials] Participants were instructed to press and hold the small button on the left side of the Actiwatch for 3 seconds each time they went to sleep and each time they woke up. Participants wore these Actiwatches to track their light data, activity data, and sleep/wake times for the duration of the study until they returned the kit.

### Salivary DLMO Collection

Before the collection, participants had access to both written instructions, visual guides, and video tutorials describing how to set up their collection space and take saliva samples. [Supplemental Material: Instructional Materials.] All study kit materials were labeled with color and symbol coded stickers to ensure participants could easily set up their necessary collection components. Twenty-four hours prior to the start of their salivary DLMO collection, participants refrained from consuming alcohol, caffeine, or any recreational drugs.^28^ On the day of the collection, participants refrained from consuming foods containing Red 40 dye or high concentrations of citric acid. Starting 6 hours before their collection, participants refrained from using nicotine or tobacco products.^28^ Prior to starting their DLMO, participants verified through the study portal that they had adhered to each of these DLMO preparation steps.

During their scheduled DLMO collection, participants collected nine samples in dim-light (<10 lux) hourly, starting 6 hours before habitual bedtime and ending 2 hours after habitual bedtime. To ease setup of the dim-light environment participants were provided with painters’ tape, contractor-grade garbage bags, aluminum foil, a light meter, and 18 tea lights which emit less than 10 lux total. Participants received automated reminder texts 45 minutes prior to the start of their DLMO and 5 minutes before each sample collection. A member of the study team was always available by text, phone call, or email to answer any questions from the participants. Participants could use electronic devices like smartphones or laptops during the collection as long as the devices were on the lowest brightness setting, set to night mode to ensure low levels of blue light emission, and at least 24 inches from their face. After participants completed their DLMO salivary sample collection, they stored their samples in their home freezers using provided freezer bags. Study data collection ended once the participant collected two home DLMO sample collections and wore their Actiwatch for at least 4 weeks. Participants shipped their kits back to the study team, including both the Actiwatch and the frozen samples from each of the DLMO collections in the provided insulated cooler box with frozen saliva samples in an additional insulated mailer package with cooler packs. [Supplemental Material: At-Home Kit Contents]

### Questionnaires

Participants completed four sets of questionnaires through the study website. Upon study enrollment, participants could complete a Health and Lifestyle Questionnaire (23 items) and a Morning-Evening Questionnaire (19 items) at their convenience during the study duration.^29^ To increase compliance for the DLMO collection instructions, participants completed a Pre-Collection Attestation directly before and a Post-Collection Attestation directly after their DLMOs. The Pre-Collection Attestation asked whether participants had abstained from substances that would interfere with melatonin production such as nicotine, caffeine, and alcohol and whether they had arranged their collection space according to the instructions. The Post-Collection Attestation asked whether the participant had collected each sample according to the provided instructions and had stored them correctly in the freezer. After they had completed their second DLMO, participants completed a Post-DLMO Questionnaire (23 items) about their experiences in the study. [Supplemental Materials: Questionnaires]

### Data Analysis

Upon receipt of the kit, each DLMO sample was inspected and qualitative temperature data was recorded (i.e. “completely frozen,” “partially frozen,” or “cool to the touch.”) Data from the Actiwatch and time-tracker bottle caps were accessed using Philips Actiware 6 Program (https://www.usa.philips.com/healthcare/sites/actigraphy/solutions/actiware) and MEMS Adherence Software (https://v4-app.aardexgroup.com/), respectively. We froze saliva samples immediately upon receipt at -80°C before shipping them to SolidPhase, Inc. (Portland, ME) for melatonin assay using the Novolytix Direct Saliva Melatonin RIA kit (ALPCO Diagnostics, Windham, NH). Intra- and inter-assay CV were 7.9% and 9.8%, respectively. DLMO time was calculated by the study team using the Hockeystick method.^30^ At the conclusion of their participation, each participant received an easy-to-read custom report that outlined their DLMO results, MEQ scores, and sleep logging data. [Supplemental Material: Participant Report Template]

A series of compliance measures were used to ensure that participants stayed on track during the study and collected valid samples: completion of both Pre- and Post-Collection Attestations, consistency of light levels below 50 lux during the collection periods, and sample collection times within 5 minutes of the scheduled collection. DLMO collection samples were considered valid only if a participant met each compliance measurement. [Figure: DLMO Compliance Measures]

## Results

Our study sample included ten individuals. Participants ranged in age from 27 to 63 years old, and the average age of the sample was 38.2 ± 11.7 years. Participants identified as female (n=7), neutrois (n=1), transgender (n=1), and male (n=1). Of the ten participants, 7 identified as “White,” and 3 identified as “Asian.” [Table: Demographics] Of the 10 participants, 1 participant had diagnosed ASWPD (MEQ score = 61), 4 participants had diagnosed DSWPD (average MEQ score = 34),, 5 participants had no CRSWD diagnosis (average MEQ score = 33.8). Participants without a CRSWD diagnosis were included to test study protocols in participants both with and without familiarity of circadian disorder diagnoses and diagnostic procedures.

Average duration of study involvement, from consent to study completion, was 42.4 days. Participants sleep logged for an average of 40.2 days and missed only 2.2 days of sleep logging on average. Five participants (50% of participants) never missed a day of sleep logging.

An advertisement on MassGeneralBrigham’s Rally site had 86 participants express interest in our study, and 30 ultimately filled out the online screener questionnaire. Twenty participants enrolled but failed to complete any study procedures, resulting in an attrition rate of 67%. Typically, enrollees would sign up and either not begin sleep logging or only sleep log for only a few days before withdrawing. Of the 20 participants who enrolled and then dropped out, they logged their sleep for an average of 2.4 days before dropping out, and 13 never began sleep logging at all. For this reason, we waited until participants had logged their sleep online for at least five days and scheduled their first DLMO with us before sending a kit.

All participants completed 2 DLMOs, each a week apart, collecting a total of 180 saliva samples across the study. Participants used our provided written and video instructions to set up their collection spaces with no study team oversight; only one participant contacted the study team with a question during their DLMO. The participants self-reported 100% compliance with pre- and post-collection protocols, as measured by the Pre- and Post-Collection Attestations.

The Actiwatch recorded light data in 30 second epochs, and this data allowed us to track whether light levels were sufficiently low during collections. Using thresholds from previous at-home DLMO studies, we considered an individual DLMO sample “compliant” if all light levels within 30 minutes of a sample collection were below 50 lux.^20^ 1 participant didn’t wear their Actiwatch during their second DLMO due to a chronic pain condition, so we had Actiwatch data for 19 DLMOs. The majority of DLMO collections were 100% compliant with light levels (<50 lux) throughout the entire collection (10/19). Of the participants who recorded high light levels (>50 lux), 7/9 of these instances occurred in the 30 minutes prior to the first scheduled collection, likely when participants were still setting up their collection space. [Table 4: DLMO Compliance Measures] One participant had numerous periods of high recorded light levels (>50 lux) throughout both of their DLMO collections (4 periods in DLMO 1, 3 periods in DLMO 2). This participant reported these exposures occurred from daylight through a window while using the restroom. They attested to wearing blue light blocking glasses during these periods, which were specifically included in the kits to mitigate the effects of high light exposure any time participants might be exposed to higher light levels (e.g., when using the bathroom). Of note, a rise in melatonin was not detected in either DLMO from this participant, indicating potential melatonin suppression.

To objectively validate the timing of sample collection, salivettes for each DLMO sample collection were retrieved from a bottle with a time stamped lid (MEMs device) which recorded the time the lid was opened. Of 180 total samples collected across all DLMO collections throughout the study, there were 7 instances (3.9% of all samples) where the timing of a saliva sample was not recorded, likely due to incomplete closure of a MEMs cap. One participant did not use the correct cap for their second DLMO, so none of the 9 sample times were recorded. Overall, 147 (82%) samples were collected within 5 minutes of the scheduled collection time. The MEMs cap timing allowed us to objectively verify sample timing and utilize accurate collection time information for DLMO calculations. [Figure: DLMO Compliance Measures] [Table: DLMO Compliance Measures]

Using salivary melatonin data, we calculated DLMO times. For 8/10 participants we were able to calculate a DLMO for at least one collection. For 6/10 participants we were able to calculate a DLMO for both DLMO collections. DLMO times were on average 3 hours and 18 minutes earlier than self-reported sleep times (DSWPD: 12:04:30 AM, ASWPD: N/A, controls: 9:55:37 PM.) [Figure: example DLMO plot] Among the 6 participants for whom we calculated two separate DLMO times, DLMOs 1 and 2 were 96% correlated (p<0.0005.) [Figure: Dlmo Correlation]

To assess participant confidence using the provided study materials (portal based, printed, and video content) and DLMO collection procedures, participants answered a Post-DLMO Questionnaire (n=23 Questions.) [Supplemental Material: Post-DLMO Questionnaire] Based on responses to the Post-DLMO Questionnaire, 70% of participants felt confident completing both DLMOs with just the written and video instructions, 100% of participants felt confident completing both DLMOs with instructions and access to a member of the study team to answer questions during the collections, although only 3 participants asked any questions during either collection. Participant confidence in performing a DLMO collection rose from DLMO collection 1 (70% feeling confident) to DLMO collection 2 (90% feeling confident). [Table: PDQ Results]

In the Post-DLMO Questionnaire, we identified challenges individual participants faced throughout the study to inform future longitudinal Circadia study. When asked about the most difficult part of the study, participants cited the length of DLMOs and/or difficulty staying awake under dim light conditions (n=5), difficulty remembering to click the event marker button on their Actiwatch at sleep and wake (n=3), difficulty of setting up DLMO space (n=2), and difficulty staying awake past their typical bedtime (n=1), suggesting future points of study protocol modification.

## Discussion

The Circadia Pilot study is one of the first studies to measure a home-based, entirely self-directed DLMO. Our study is unique due to the fact that there are absolutely no in person visits required and only communication with the study team if the individual needs it. It is also one of the first home studies to perform repeat DLMOs in healthy individuals and CRSWD patients in the same study. While previous studies have required in-person instruction from study staff, participants in this study were able to comply with study protocols using instructional videos and written guides, with optional access to study staff. We made circadian phase assessment more accessible by making the assay completely home-based and self-directed with a low-cost kit containing all necessary components to complete the assessment. This study, with both CRSWDs patients (N=5) and controls (N=5), found the at-home kits to be reliable and produce similar results to those of a supervised, in lab test. Using the kits, we verified CRSWD diagnoses and used objective measures to verify testing procedure was followed. With the kit materials, instructional videos, 24-hour access to study materials, and study staff on call to help participants during sample collections, our protocol mimics those of a supervised, in hospital and/or laboratory study without the barriers of cost, wait time, and travel restrictions.

While overseeing DLMOs remotely, we are able to objectively measure participant compliance to best practices for each individual saliva sample using Actiwatch and MEMs data to capture activity, light, and sample collection time data objectively. Most instances of non-compliance with light level measures occurred prior to participants’ first DLMO sample collection, indicating a difficulty remaining in low light while participants were still setting up their collection space prior to their first sample collection. This should be remedied in future studies by instructing participants to set up and enter their study area 30 minutes prior to first sample collection.

There were two cases for which we could not calculate a DLMO time using the data acquired. The challenge is to distinguish errors in sample collection from biologically complicated cases. In the first case, the participant was the only individual diagnosed with ASWPD. This individual’s salivary melatonin levels consistently measured low (< 2.5 pg/mL) throughout the collection period rendering calculation of a DLMO time using any published methods impossible. In rare cases of low salivary melatonin producers, it may not be feasible to use salivary melatonin data for DLMO time.^31^ Actiwatch data collected during this participant’s DLMO collections, however, showed 4 instances of non-compliant (>50 lux) light levels during DLMO 1 and 3 instances during DLMO 2, potentially resulting in melatonin suppression. While we cannot rule out biological low production of melatonin, our objective compliance measures point toward a need for repeat DLMO testing and increasing the sample size in order to increase the power for determining accurate DLMO time.

In the second case, the participant, who had been diagnosed with DSWPD, displayed highly inconsistent sleep times and a history of DLMO assays that failed to attain a circadian phase profile. In outlier cases like these, a salivary DLMO is likely insufficient to capture circadian phase, and a more sensitive 24-hour urine test may be called for.^32^ As we were unable to identify a DLMO in either collection from this participant, we suspect an underlying biological variability for this participant and would suggest 24-hr urinary melatonin collection to obtain an accurate DLMO profile. These cases are likely representative of limitations of salivary melatonin-based circadian phase assessments in general.^31, 32^

A key feature of the study design as a direct-to-participant study was its ease of enrollment. Consistent with a direct-to-participant approach, our recruitment and enrollment procedure were designed so that participants could enroll online with as few barriers to entry as possible. As a byproduct of this ease of enrollment, we anticipated a high attrition rate early in study participation, and we found this to be true among participants within the first few days after enrollment. This study design enabled participants to familiarize themselves with the study procedures and get a realistic idea of study burden before deciding to fully commit. Participants who lost interest or decided they could not commit were able to easily unenroll without significant wasted time or resources spent either by participants or the study team.

Our results suggest that autonomous, self-directed, at-home DLMOs are accurate and feasible. One current limitation of the study is the lack of temperature data for samples in transit. Since temperature can affect the stability of salivary melatonin,^33^ future studies will utilize temperature sensors in the at-home study kit. Another limitation of the study was the lack of individuals with a ASWPD diagnosis (N=1). We were unsuccessful at obtaining DLMO times for them and as our only ASWPD participant, we were unsuccessful at making conclusions of our kit on this patient population. Importantly, our methods are generalizable, accessible, and relatively inexpensive. As circadian rhythms have been implicated in a range of health conditions,^6^ circadian phase data can be a useful metric for studies beyond CRSWDs. We hope our at-home self-directed DLMO design can serve as a framework to implement circadian phase assessment in many other contexts.

## Supporting information

Table 1. Demographics

Table 2. Kit Contents

Table 3. PDQ Results Summary

Table 4. DLMO Compliance Percentages

Figure 1. DLMO Compliance Measures

Figure 2. Sample DLMO Plot

Figure 3. DLMO Correlation

Circadia Study Participant Template

## Data Availability

All data produced in the present study are available upon reasonable request to the authors.

## Acknowledgments

This research and Prof Jacqueline M. Lane was supported by grants from the Massachusetts General Hospital Mc Cancer Center SPARC Award, the Massachusetts General Hospital Claflin Distinguished Scholar Award, and NIH Grant K01HL136884. We thank the Broad Institute Data Science Platform for guidance on direct-to-patient study and portal design.

## Author Contributions

*Study conceptualisation and design:* JML, MBW, OJA, SR, JW.; *Analysis:* JC, GB, JL, JML.; *Overseeing project:* JML, JL, GB.; *Writing and editing:* JML, JL, GB.; *Critical revisions of the manuscript:* GB, JL, JC, MBW, OJA, SR, JW, JML.

## Disclosure Statements

### Financial statement

None.

### Conflicts of interest statement

The funders had no role in study design, data collection and analysis, decision to publish, or preparation of the manuscript.

## Notes

### Competing Interest Statement

The authors have declared no competing interest.

### Funding Statement

This work was supported by NIH grants K01HL136884 (JML), and The Massachusetts General Hospital McCance SPARC Award, Claflin Award, and from the National Center for Advancing Translational Science. Center (P30- DK040561) Nutrition Obesity Research Center at Harvard.
The authors have no competing financial interests to declare.

### Author Declarations

IRB of MassGeneralBrigham gave ethical approval for this work.

