## Supplementary figures and images for "Self-Directed Home-Based Dim-Light Melatonin Onset Collection: The Circadia Pilot Study"

### Circadia Study Participant Template

#### Slide 1
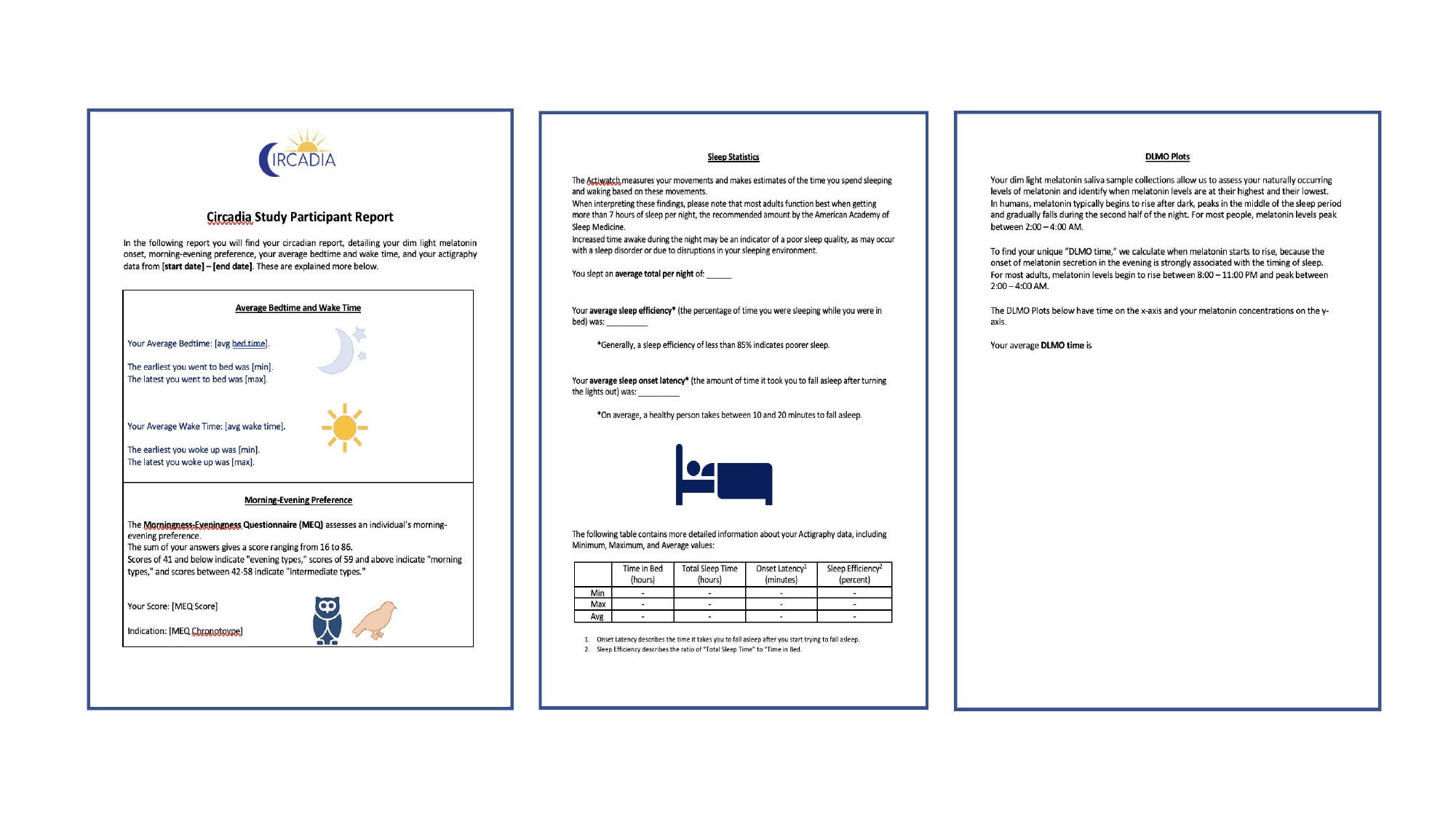

### Figure 1. DLMO Compliance Measures

#### Slide 1
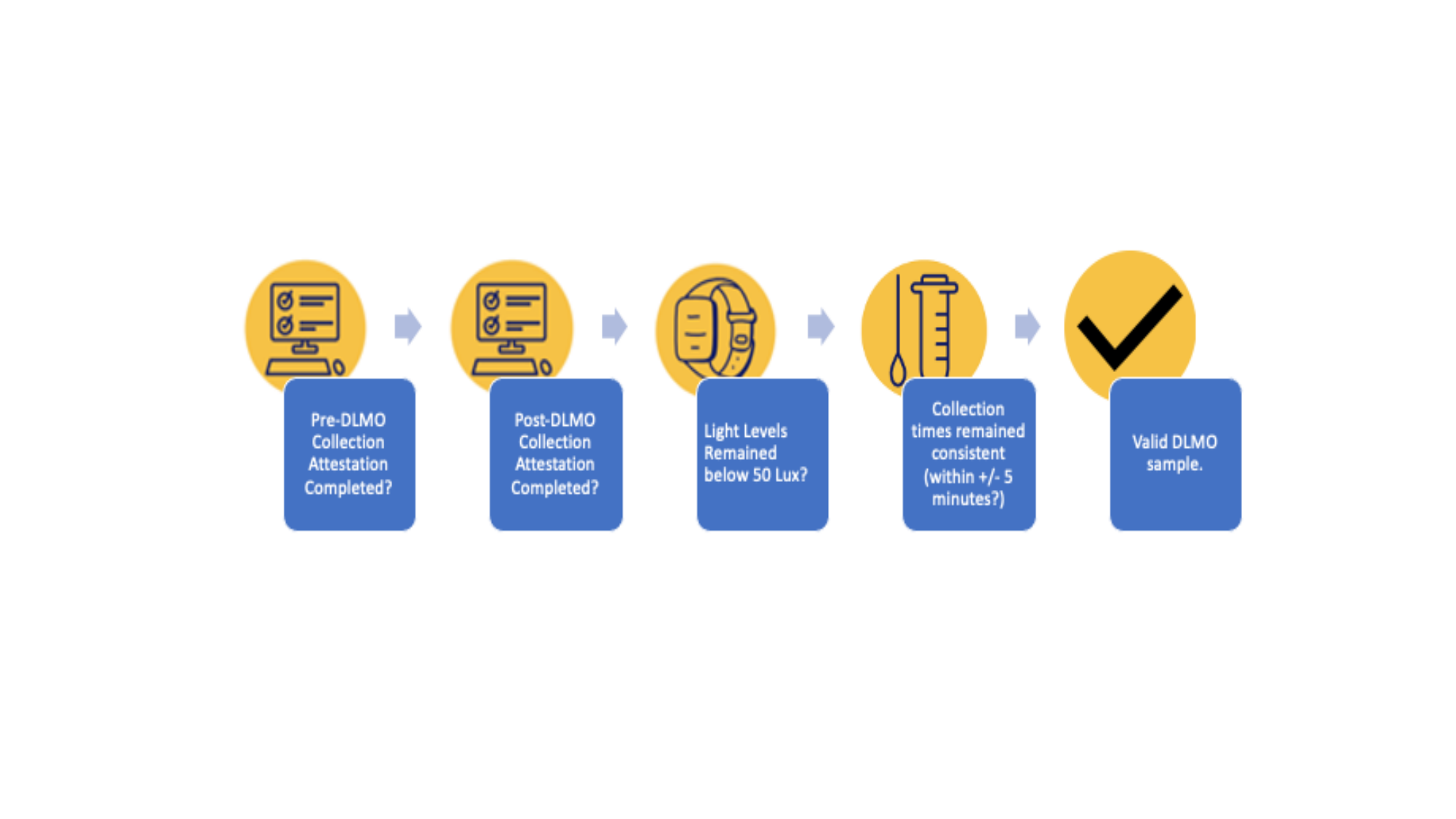

### Figure 2. Sample DLMO Plot

#### Slide 1
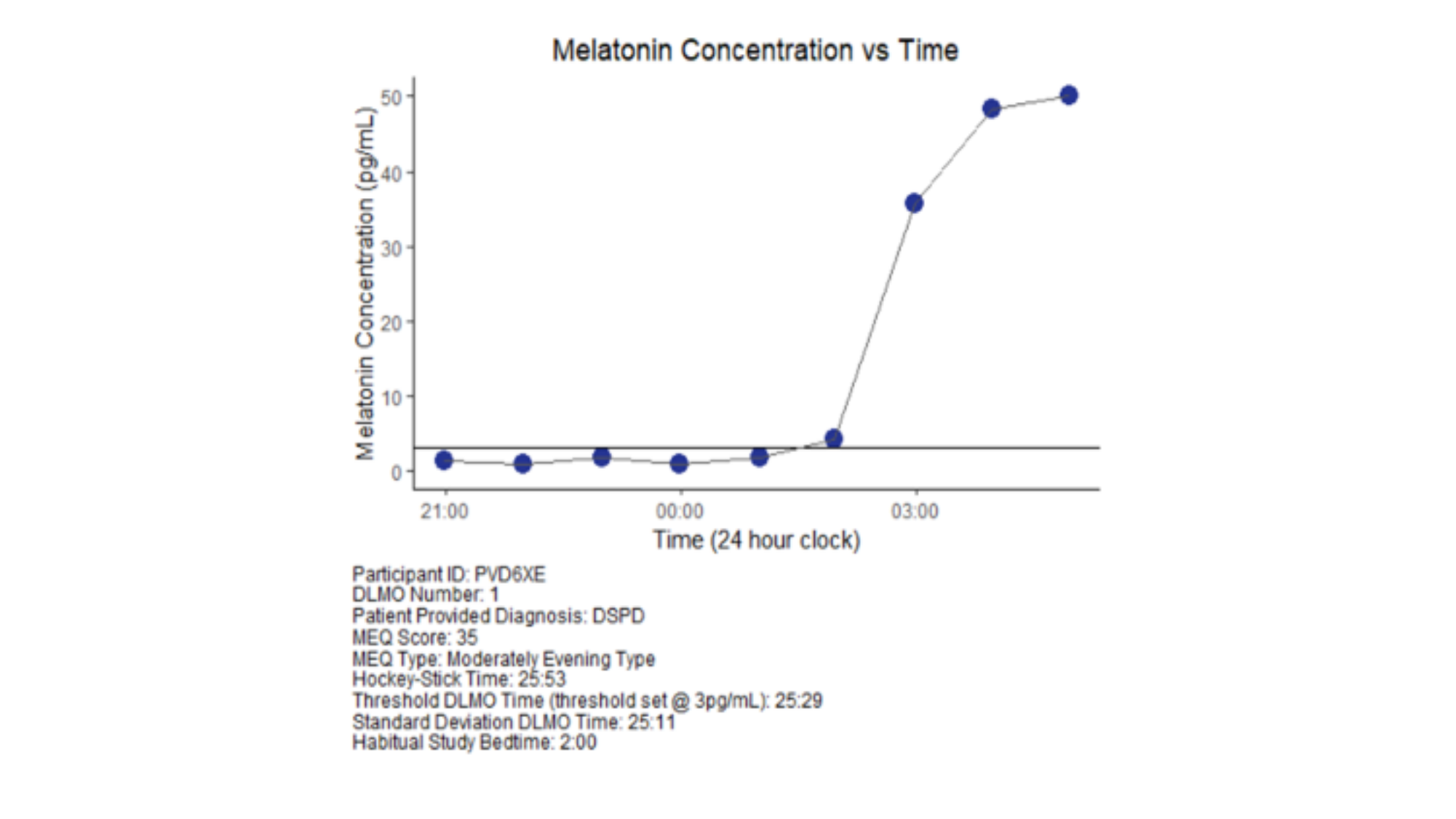

### Figure 3. DLMO Correlation

#### Slide 1
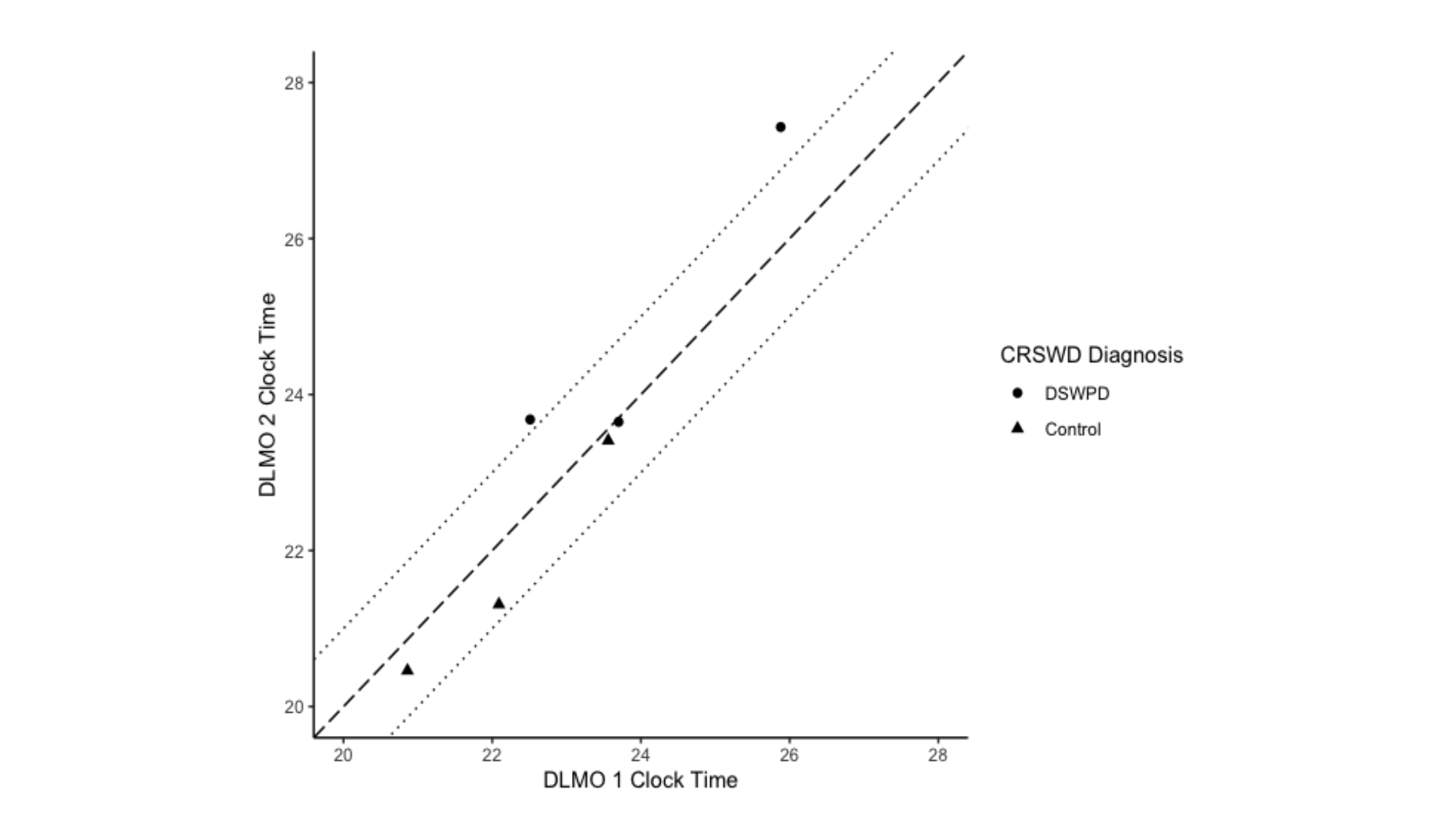

### Table 1. Demographics

#### Slide 1
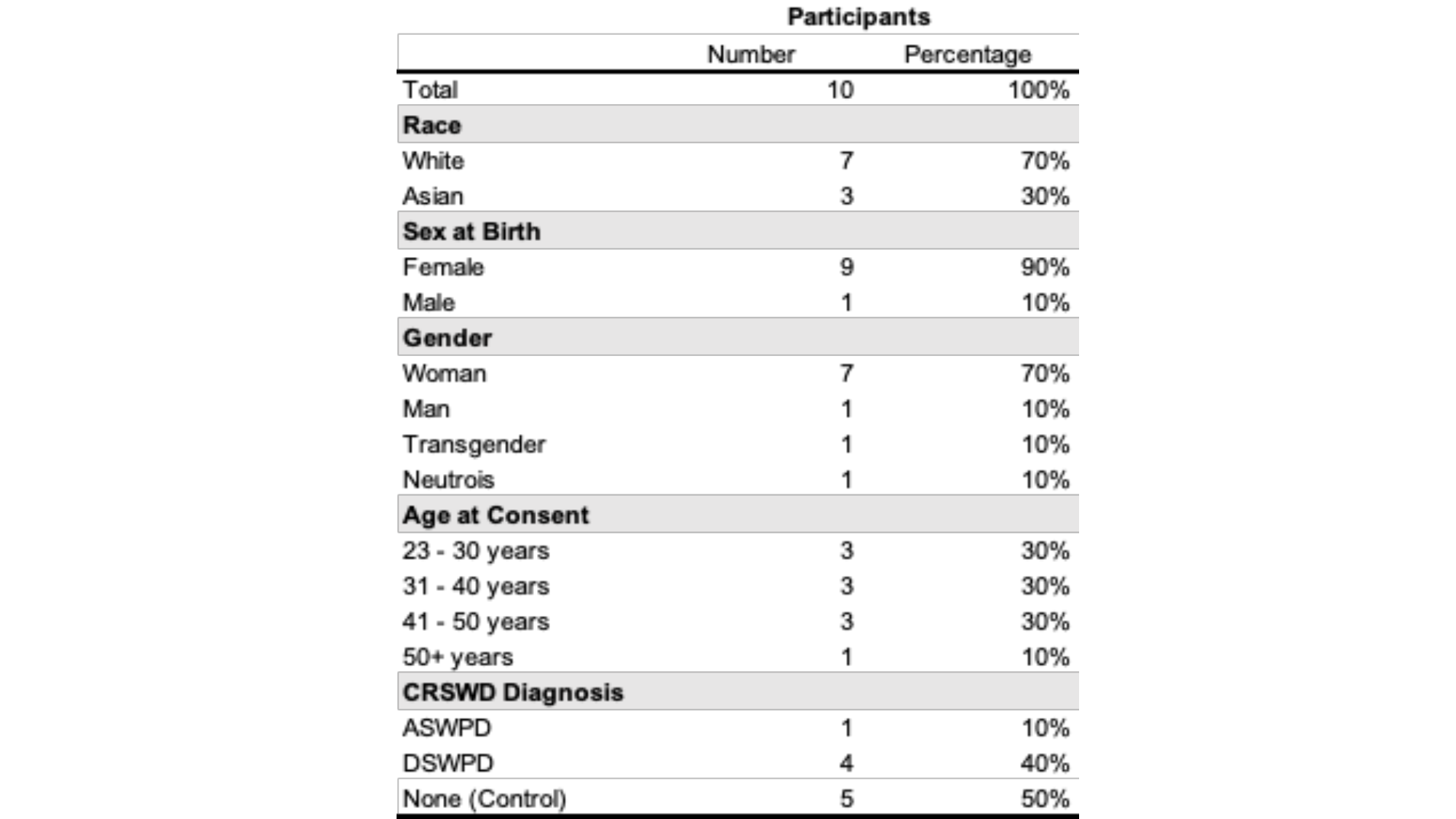

### Table 2. Kit Contents

#### Slide 1
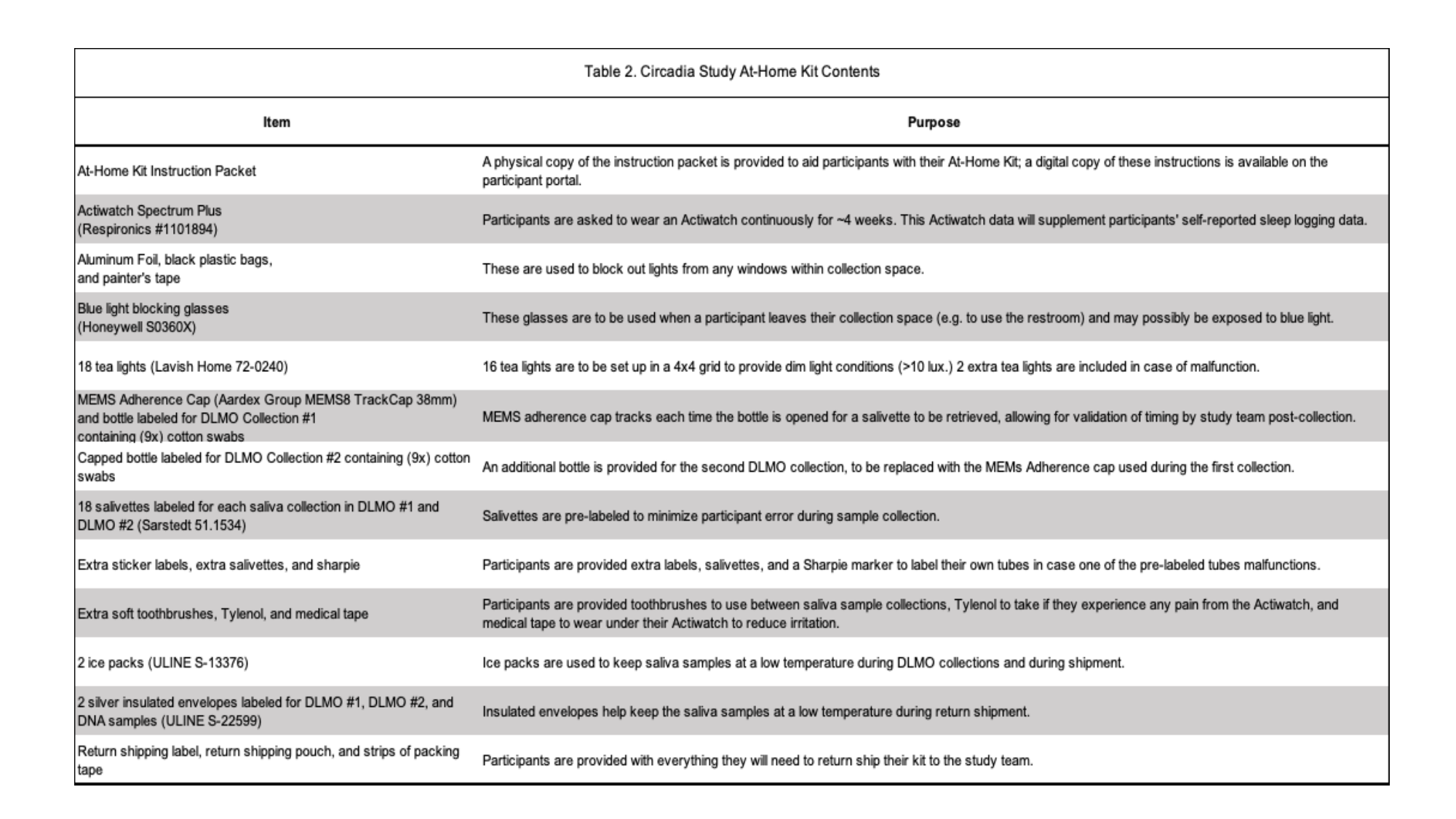

### Table 3. PDQ Results Summary

#### Slide 1
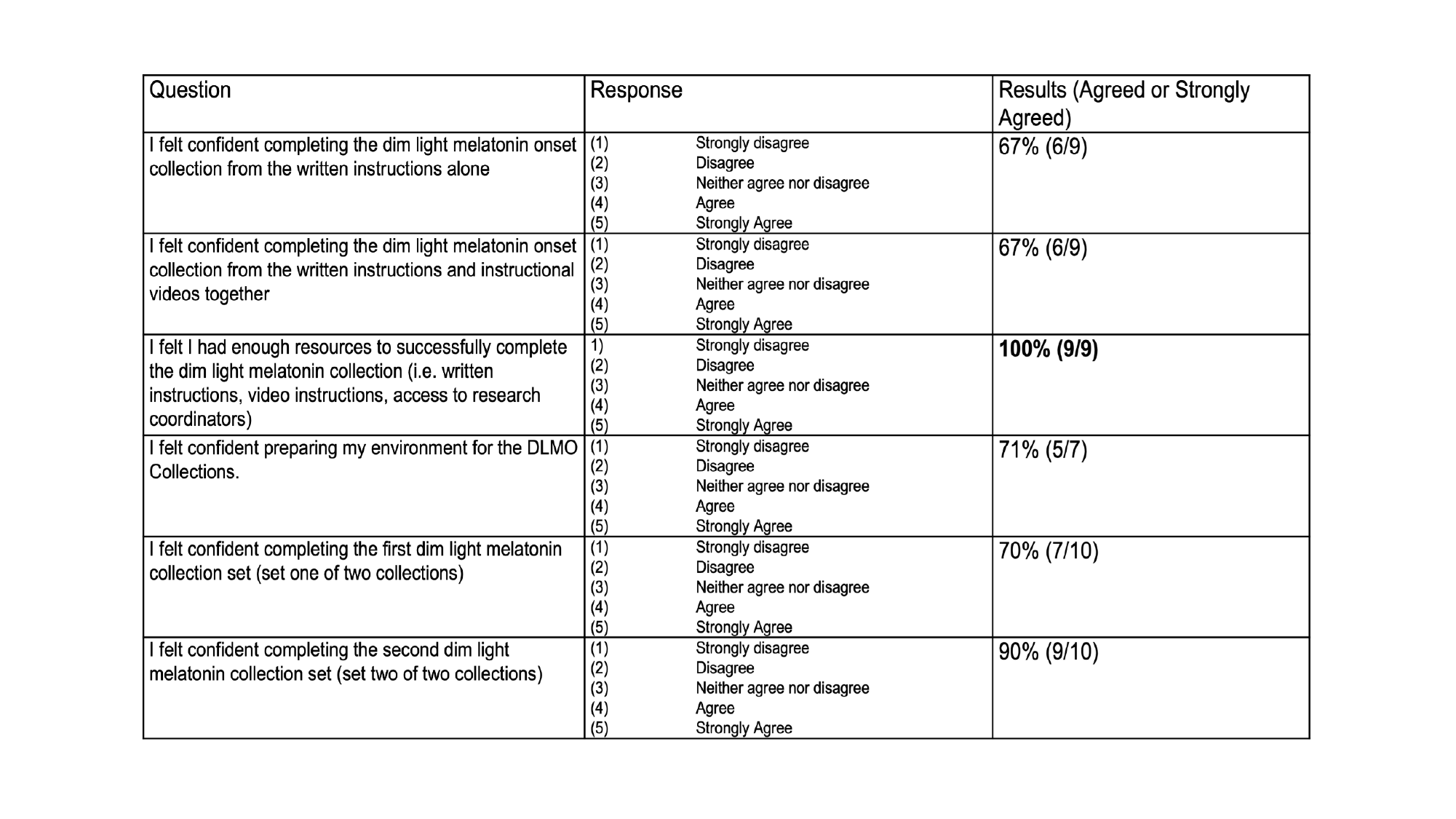

### Table 4. DLMO Compliance Percentages

#### Slide 1
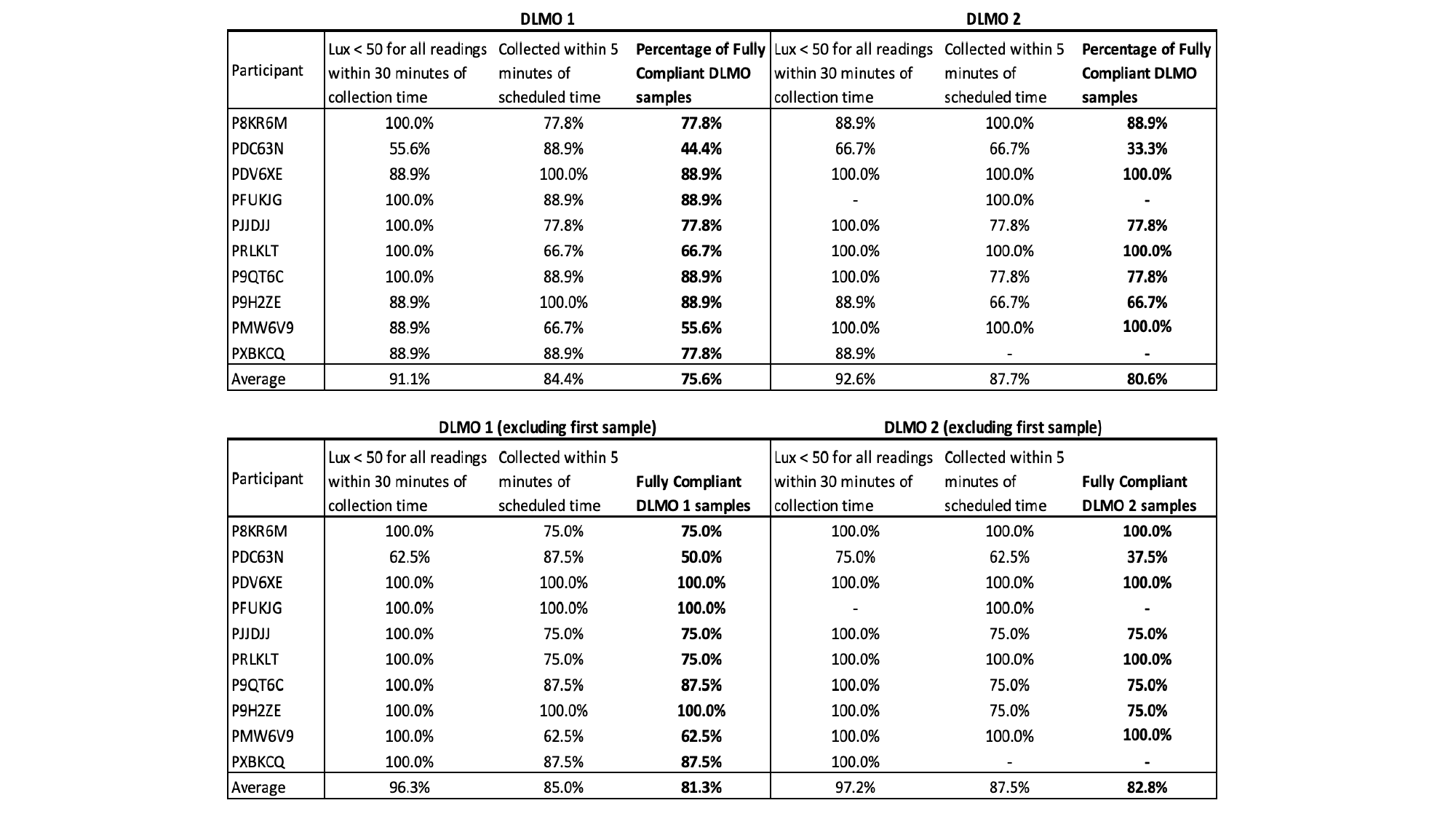
